# 15-month follow-up of anti-spike receptor binding domain (RBD) SARS-CoV-2 antibodies among healthcare workers in Boston, MA

**DOI:** 10.1101/2022.03.18.22272553

**Authors:** Maura C. Dodge, Manisha Cole, Elizabeth R. Duffy, Martha M. Werler, Yachana Kataria

**Author notes:** Corresponding author**Address for correspondence:** Yachana Kataria, Medical Director of Clinical Chemistry, Boston Medical Center, Department of Pathology and Laboratory Medicine, 670 Albany St, Boston, MA 02118.

## Abstract

Over 15-months we found that anti-spike RBD SARS-CoV-2 antibody concentrations follow different trends with combinations and permutations of COVID-19 infection and vaccination among healthcare workers in Boston, MA. A majority of HCWs remain well above the positivity threshold for anti-spike RBD IgG antibodies for at least 9 months following vaccination regardless of infection history. Of interest, those with COVID-19 infection before vaccination had significantly higher median serum antibody concentrations in comparison to HCWs with no prior infection at each follow-up timepoint. These findings further support what is known regarding the decline in serum antibody concentrations following natural infection and vaccination, adding knowledge of serum antibodies up to 15 months post infection and 11 months post vaccination.

**Importance:** Boston Medical Center (BMC) is a safety net hospital in Boston and from the initial wave of COVID-19 there has been overwhelming concern about the exposure of healthcare workers to SARS-CoV-2. We conceived a longitudinal study to assess virus exposure and trends in SARS-CoV-2 antibodies amongst healthcare workers at BMC over 15 months. We have followed HCWs through three waves of COVID-19, including the Delta variant wave from June through mid-December 2021, assessing anti-spike receptor binding domain IgG, anti-nucleocapsid IgG, and anti-spike IgM at approximately three-month intervals. Current literature largely describes antibody durability six months post vaccination. These data add to the literature by describing antibody durability and trend differences according to infection history and vaccination status. These longitudinal data contribute to a greater understanding of the ongoing COVID-19 pandemic and can help inform future research and public health decision-making regarding vaccine uptake, breakthrough infections, and overall pandemic response.

## Introduction

Positive antibody response trends following vaccinations and/or natural infection with COVID-19 have been identified and reported (1,2). Together, declining antibody concentrations observed among individuals in convalescence and those vaccinated who initially exhibited strong anti-spike IgG responses point to potentially universal waning in long-term protection (2,3). It remains unclear how antibody levels change over time beyond 6 months after vaccination and how these levels may correlate with protection (2).

It is critical to understand how combinations of natural and vaccine-derived antibodies protect healthcare workers (HCWs), an important population at an increased risk of exposure to COVID-19 (4,5). With the advent of booster vaccines targeting the wild type virus sequences and the occurrence of breakthrough infections from variants, observing the evolution of anti-COVID-19 antibodies among HCWs *with* and *without* a history of infection is critical to informing public health decisions. We followed serum concentrations of anti-spike receptor binding domain (RBD) SARS-CoV-2 IgG within a cohort of HCWs at Boston Medical Center (BMC), Boston, Ma over 15 months in 3-month intervals, observing trends by vaccination status and infection over time.

## Methods

At baseline in July 2020, 527 Boston Medical Center HCWs provided serum samples and survey data on demographic and health-related information pertaining to the previous three months including symptoms, comorbidities, and RT-PCR testing history. Participants returned in December 2020, March 2021, June 2021, and September 2021 to provide updated survey data and blood samples. Self-reported COVID-19 vaccine status and RT-PCR test results were confirmed where possible using electronic medical records (EMR). Questionnaires were collected and managed in REDCap electronic data capture tools hosted at Boston University, CTSI 1UL1TR001430 (6,7). This study was approved by the Institutional Review Board at BMC (8).

Samples from all timepoints were analyzed for SARS-CoV-2 anti-spike-1 RBD IgG II (quantitative) and anti-nucleoprotein IgG (qualitative) in the clinical pathology laboratory at BMC using the Abbott Architect i2000sr, using IgG II (quantitative) and IgG (qualitative) assays, respectively (Abbott Laboratories, Abbott Park, IL) following manufacturer’s specifications. Samples with a concentration of ≥50 AU/mL were positive for anti-s IgG (quant) and values were subsequently log_2_ transformed for visualization. Anti-n IgG (qual) values were interpreted as positive with an index value ≥1.4 (8).

Seropositivity and infection history were determined by RT-PCR results and antibody status. Specifically, both RT-PCR results and anti-n IgG (qual) results were used to assist in identifying cases of natural COVID-19 infection throughout follow-up, whereas anti-s IgG (quant) results were used to analyze serology trends within this cohort. The Abbott anti-s IgG (quant) assay became available after the initial time point and bio-banked serum samples were used to examine serum antibody concentrations for the prior timepoints. Participants were grouped according to their vaccination and infection status in September 2021 into (1) infected before vaccination, (2) infected after vaccination, (3) vaccinated with no history of infection, (4) unvaccinated with no history of infection, or (5) unvaccinated with a history of infection.

Statistical analyses were run using Statistical Analysis Software version 9.4 (SAS Institute, Cary, NC). Box plots visualize each group’s median anti-s IgG concentrations with whiskers depicting the 1.5*IQR rule at each data collection timepoint. Mood’s median testing was used to compare median serum antibody concentrations between groups at all timepoints.

## Results

Of the 527 enrolled in July 2020, participants were predominantly female (77.0%) and approximately half self-reported a normal BMI (47.4%). Most identified as White (81.6%); the remaining as Asian, Black, Mixed Race, or Other (8.4%, 3.8%, 2.5%, and 2.9%, respectively). Ethnicity was reported as Hispanic/Latinx by 7.4%. Participants were largely nurses (41.2%) or physicians (31.5%).

### Unvaccinated HCWs

Unvaccinated and never positive HCWs expectantly never crossed the positivity threshold and were all lost to follow-up by June 2021 (July 2020: n=42, December 2020: n=42, and March 2021: n=3). Unvaccinated HCWs with a history of infection saw fluctuating anti-s IgG positivity corresponding to varying dates of COVID-19 positivity over the course of the study (n=17, 15, 6, 7, 4) (Table 1, Figure 1).

**Table 1.** Median and interquartile range values for anti-s receptor binding domain SARS-CoV-2 serum antibody concentrations by vaccination and natural infection group among Boston Medical Center HCWs over 15 months of follow-up

| domain SARS-CoV-2 serum antibody concentrations by vaccination and natural infection group among Boston Medical Center HCWs over 15 months of follow-up |  |  |  |  |
| --- | --- | --- | --- | --- |
| Category | Serology date | n | Median*, AU/mL | IQR |
| Infected before vaccination | July 2020 | 72 | 233.2 | 837.9 |
|  | December 2020 | 67 | 374.5 | 612.5 |
|  | March 2021 | 57 | 25534.5 | 45090.2 |
|  | June 2021 | 57 | 8342.2 | 14400.5 |
|  | September 2021 | 51 | 4463.2 | 7043.1 |
| Vaccinated with no history of infection | July 2020 | 352 | 1.3 | 2.1 |
|  | December 2020 | 330 | 1.1 | 2.3 |
|  | March 2021 | 307 | 10012.3 | 11304.7 |
|  | June 2021 | 278 | 2406.3 | 3046.8 |
|  | September 2021 | 247 | 999.3 | 1258.1 |
| Infection after vaccination | July 2020 | 11 | 1.3 | 1.2 |
|  | December 2020 | 11 | 1.3 | 1.4 |
|  | March 2021 | 11 | 8445.8 | 4599.0 |
|  | June 2021 | 10 | 1811.6 | 1434.5 |
|  | September 2021 | 8 | 989.4 | 3419.2 |
| Unvaccinated with a history of infection | July 2020 | 17 | 571.5 | 826.8 |
|  | December 2020 | 15 | 359.6 | 381.0 |
|  | March 2021 | 6 | 389.5 | 331.2 |
|  | June 2021 | 7 | 450.3 | 2592.8 |
|  | September 2021 | 4 | 11269.7 | 24631.2 |
| Unvaccinated with no history of infection | July 2020 | 42 | 1.8 | 2.4 |
|  | December 2020 | 42 | 1.4 | 1.9 |
|  | March 2021 | 3 | 1.0 | 2.3 |

\*Positivity threshold for IgG Quant assay is 50 AU/mL

**Figure 1.**
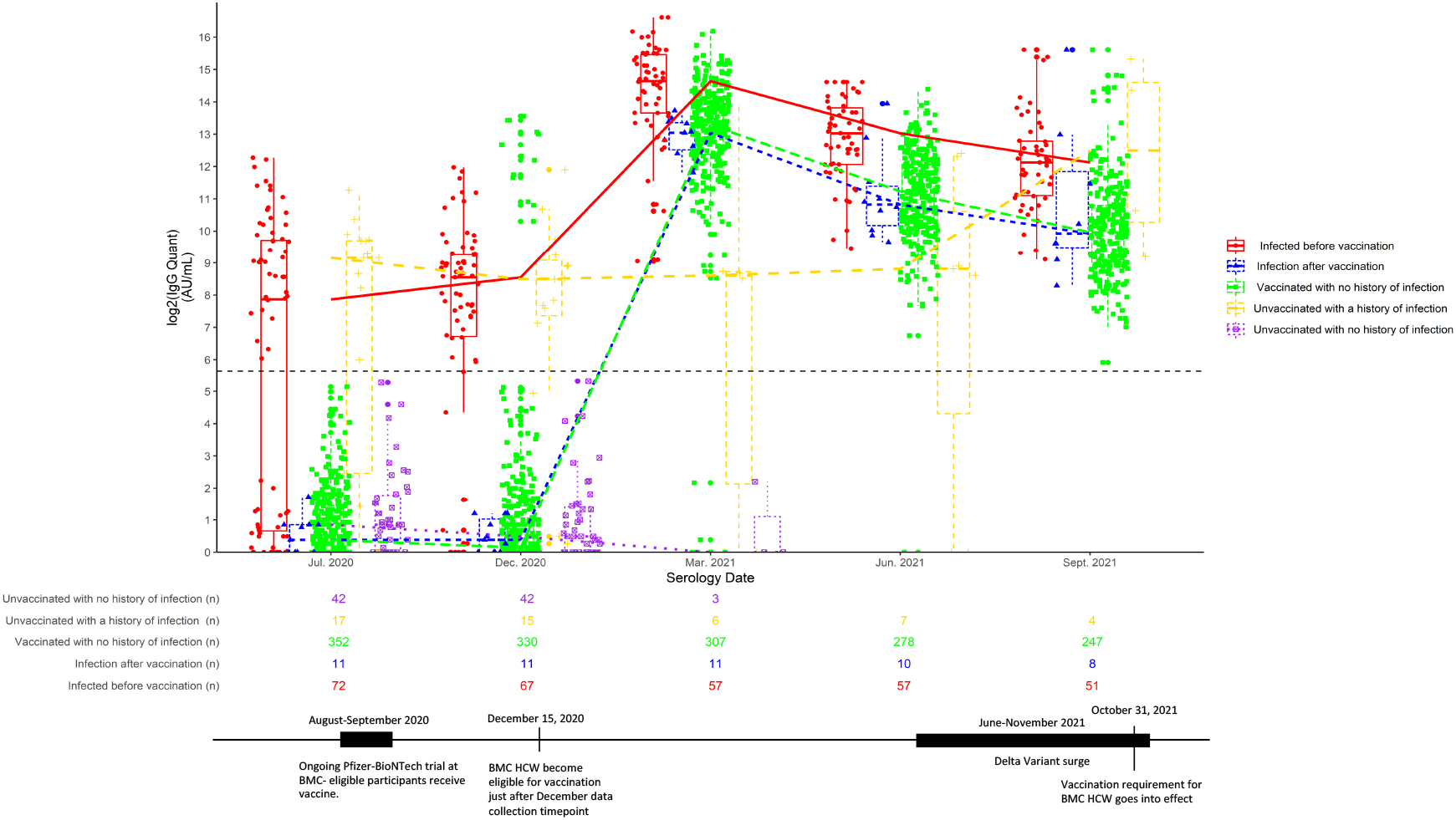
Boxplots of log_2_-transformed anti-spike receptor binding domain SARS-CoV-2 IgG antibody concentrations (AU/mL) over 15 months. The dashed line represents positivity threshold for IgG Quantitative assay (≥50 AU/mL, log2-transformed positivity threshold: 5.64 AU/mL) and the superimposed timeline describes important milestones in the COVID-19 pandemic. Box plots with whiskers represent the median and first and third quartiles extended by 1.5 times the interquartile range.

### Infected before vaccination

At baseline (n=72), median anti-s IgG concentration for those with infection prior to vaccination was 233.2 AU/mL (IQR, 837.9 AU/mL), leveled to 374.5 AU/mL (IQR, 612.5 AU/mL) in December 2020 (n=67), and dramatically increased to 25,534.5 AU/mL (IQR, 45,090.2 AU/mL) in March 2021 (n=57), concurrent with major vaccine uptake. Vaccinations were available to all BMC HCWs in mid-December 2020, with most reporting full vaccination between January and February of 2020. From March to June 2021, an approximate 66% decline in concentration to 8,342.2 AU/mL (IQR, 14,400.5 AU/mL) was observed (n=57), followed by further decline to 4,463.4 AU/mL (IQR, 7,043.1 AU/mL) in September 2021 (n=51) for an overall 82.5% decline (Table 1, Figure 1).

### Vaccinated and never infected

A majority of HCWs were vaccinated with no infection and saw peak median IgG levels in March 2021 coinciding with major vaccine uptake (10,012.3 AU/mL, IQR 11,304.7 AU/mL) (n=307). Concentrations declined almost 76% by June 2021 to 2,406.3 AU/mL (IQR, 3,046.8 AU/mL) (n=278) and continued to decline an overall 90.0% into September (999.3 AU/mL, IQR 1,258.1 AU/mL) (n=247) (Table 1, Figure 1).

Mood’s median testing confirmed that following widespread vaccination in March 2021, those who were infected before full vaccination had significantly different median anti-s RBD IgG serum concentrations (25,112.10 AU/mL) compared to vaccinated individuals without a history of infection (10,082.45 AU/mL) (p<0.0001). Additionally, in June and September 2021 the median values were significantly different with those infected before vaccination showing median concentrations of 7,588.80 AU/mL and 4,463.40 AU/mL, respectively, compared to those vaccinated with no history of infection (2,406.30 AU/mL in June and 1,002.05 AU/mL in September) (p<0.0001).

Twelve participants were enrolled in an ongoing Pfizer-BioNTech vaccine trial at BMC and were administered the vaccine in August and September 2020. We observed positive antibody levels during their next follow-up visit in December 2020 (median, 4,685.1 AU/mL). In September 2021, eleven months after vaccination, median serum concentrations in this group remained positive but had declined by 92% to 369.5 AU/mL.

### Infection after vaccination

In March 2021, antibody concentrations for those infected after at least one vaccine dose rose from below the positivity threshold to 8,445.8 AU/mL (IQR, 4,599.0 AU/mL) (n=11) before a steady decline to 1,811.6 AU/mL (IQR, 1,434.5 AU/mL) in June 2021 (n=10) and 989.4 AU/mL (IQR, 3,419.2 AU/mL) in September 2021 (n=8) (Table 1, Figure 1). From universal positivity in March 2021 to the most recent timepoint in September 2021, median antibody concentrations declined by 88.3% overall for these participants.

## Discussion

Observing longitudinal trends of SARS-CoV-2 antibodies provides insight into protection from reinfection and vaccine efficacy. HCWs with positive anti-s IgG antibody concentrations following vaccination and natural infection had variably declining antibody levels over the 15-month follow-up period. Unvaccinated individuals, a small portion of the cohort, showed positive anti-s IgG concentrations following infection before loss to follow-up. Among vaccinated participants, anti-s IgG levels declined but largely remained above the positivity threshold up to 11 months post-vaccination regardless of infection history. HCWs that participated in the Pfizer-BioNTech vaccine trial and were fully vaccinated in September 2020 all remained positive for serum anti-s IgG for 11 months, but their median antibody titers were only 8% of their initial peak levels. Collectively our data suggest that positive anti-s IgG serum antibody concentrations persist for up to 15 months but decline at variable rate with different combinations and permutations of vaccination and COVID-19 infection.

Our data is consistent with previously published data demonstrating an overall decline, but continued positivity up to 6 months post-vaccination, with some models predicting continued positivity for up to 2 years (2,4,9). Importantly, our findings show that amongst vaccinated HCWs, anti-s IgG antibodies can persist above the positivity threshold for up to 11 months. Our findings of continued seropositivity beyond that which has been published previously are important for understanding long-term kinetics and durability of SARS-CoV-2 antibodies.

Eight of the 11 individuals classified as breakthrough cases were infected during their vaccination sequence (determined through post-hoc EMR analysis). There were three true breakthrough cases of COVID-19 infection (occurring at least two weeks after full vaccination) with variable reported symptoms and IgG levels (1,038.2-8,626.0 AU/mL). Of interest, the participant with the lowest serum IgG concentration reported the most symptoms (dyspnea, runny nose, headache, and cough), suggesting that symptom severity with breakthrough infections could be influenced by the serum concentrations of antibodies. These results are in accordance with publications describing correlates of protection against symptomatic infection and asymptomatic infection showing that higher serum antibody titers were associated with a lower risk of infection and adverse outcomes from COVID-19 (10, 11). Additionally, an overall breakthrough rate of 0.92% in our high-risk population, all of whom reported mild infections, highlights vaccine efficacy in our cohort.

Currently it is unknown what titer of SARS-CoV-2 IgG levels equates to protective immunity against reinfection or influences disease severity. Further, duration of the presumed protection is also unknown but the potential for long-term duration of protective immunity is not unfounded. Studies suggest that neutralizing antibodies could serve as a correlate of protection for vaccines against SARS-CoV-2 (10). We were unable to measure neutralizing antibodies as these assays involve live viral culture and require biosafety level 3 containment facilities. However, others have shown that binding and neutralizing antibody titers are thought to be correlated with each other (10-14). Specifically, the anti-s RBD IgG assay used here has been highly correlated with neutralizing antibodies elicited by vaccination strengthening our use of this quantitative measure of immunity in our cohort (12).

Booster doses were approved for HCWs in the US on September 22, 2021, after our most recent serology time point, and accordingly we have not yet seen any appreciable trends from this cohort. Of note, one participant had received a booster vaccine dose prior to the September 2021 timepoint and elicited peak antibody concentration comparable to HCWs infected prior to vaccination (20, 205.1 AU/mL) suggesting that an additional vaccine dose can elicit a similar response to infection before vaccination. Ultimately, observing antibody kinetics following these doses and characterization of breakthrough infections will be important for monitoring reinfection rates and developing strategies for pandemic response. It has been shown that even individuals with positive binding and neutralizing antibody titers continue to be at risk for symptomatic and asymptomatic COVID-19 infection, as seen with SARS-CoV-2 Omicron variant, highlighting that with continued virus mutation, new variants can effectively evade this acquired immunity (15, 16, 17).

Long term immunity can be conferred by humoral and cell-mediated immune responses. The spike glycoprotein contains the receptor-binding domain, rendering it the main target for neutralizing antibodies and vaccines. This study did not assess the complete immune profile of the participants, limiting our conclusions about long term immunity that can be imparted by the interplay between humoral and cellular immune responses. Our results show that anti-s IgG antibodies, while variable amongst participants, remain above positivity thresholds up to 15 months following infection and 11 months after vaccination. Collectively, our results indicate that over 15 months of longitudinal follow-up, antibody durability and serum concentration trends resulting from vaccines and infection continue to vary by vaccination and infection status.

We limited misclassification bias and strengthened our findings by confirming self-reported vaccination status and RT-PCR results in EMRs, when possible, and by following our population for 15 months beginning early in the pandemic (July 2020) and maintaining 62% of the initial participants through to September 2021. However, we are unable to generalize to the greater public because our population is largely young and healthy. We plan to follow this cohort for an additional timepoint to gather further insight into antibody levels, particularly with the uptake of booster vaccine doses among HCWs. Given the rise of new variants of concern (specifically the Omicron variant), it is vital to determine correlates of protection from COVID-19.

In conclusion, we report waning IgG levels among HCWs stratified by infection history and vaccination status over 15 months with significantly higher antibody concentrations in participants with COVID-19 infection prior to vaccination. Our findings provide new longitudinal data that detectable antibody levels persist for 11 months post-vaccination and support the continued surveillance of SARS-CoV-2 antibody kinetics and determining correlates of protection.

## Data Availability

All data produced in the present study are available upon reasonable request to the corresponding author.

## Acknowledgements

We acknowledge the clinical chemistry, phlebotomy, and central receiving staff in the Department of Pathology and Laboratory Medicine at Boston Medical Center for their work to help accomplish this study.

This research was supported, in part, by BMC Development Philanthropy Funds for COVID-19 research and Abbott Core Diagnostics.

## References

1. Post, N., Eddy, D., Huntley C., van Schalkwyk M., Shrotri M., Leeman, D., Rigby, S., Williams, S.V., Bermingham, W.H., Kellam, P., Maher, J., Shields, A.M., Amirthalingam, G., Peacock, S.J., and Ismail, S.A. Antibody response to SARS-CoV-2 infection in humans: A systemic review. PLoS ONE. 2020 Dec; 15(12). Available from: https://doi.org/10.1371/journal.pone.0244126.

2. Naaber, P., Tserel, L., Kangro, K., Sepp, E., Jurjenson, V., Adamson, A., Haljasmagi, L., Rumm, A.P., Maruste, R., Karner, J., Gerhold, J.M., Planken, A., Ustav, M., Kisand, K., and Peterson, P. Dynamics of antibody response to BNT162b2 vaccine after six months: a longitudinal prospective study. The Lancet Regional Health-Europe. 2021 Nov; 10(100208). Available from: https://doi.org/10.1016/j.lanepe.2021.100208.

3. Lau, E., Hui, D., Tsang, O., Chan, W.H., Kwan, M., Chiu, S., Cheng, S., Ko, R., Li, J., Chaothai, S., Tsang, C., Poon, L., and Peiris, M. Long-term persistence of SARS-CoV-2 neutralizing antibody responses after infection and estimates of the duration of protection. EClinical Medicine, The Lancet. 2021 Nov; 41(101174). Available from: https://doi.org/10.1016/j.eclinm.2021.101174.

4. Hughes M.M., Groenewald, M.R., Lessem S.E., Xu, K., Ussery, E.N., Wiegand R.E., Qin, X., Do, T., Thomas, D., Tsai, S., Davidson, A., Latash, J., Eckel, S., Collins, J., Ojo, M., McHugh, L., Li, W., Chen, J., Chan, J., Wortham, J.M., Reagan-Steiner, S., Lee, J.T., Reddy, S.C., Kuhar, D.T., Burrer, S.L., Stuckey, M.J. Update: Characteristics of health care personnel with COVID-19 – United States, February 12-July 16, 2020. MMWR Morb Mortal Wkly Rep. 2020 Sept; 69(38). Available from: https://doi.org/10.15585/mmwr.mm6938a3.

5. Nguyen L.H., Drew D.A. Graham, M.S., Joshi, A.D., Guo, C.G., Ma, W., Mehta, R.S., Warner, E.T., Sikavi, D.R., Lo, C.H., Kwon, S., Song, M., Mucci, L.A., Stampfer, M.J., Willet, W.C., Eliassen, A.H., Hart, J.E., Chavarro, J.E., Rich-Edwards, J.W., Davies, R., Capdevila, J., Lee, K.A., Lochlainn, M.N., Varavsky, T., Sudre, C.H., Cardoso, M.J., Wolf, J., Spector, T.D., Ourselin, S., Steves, C.J., and Chan, A.T. Risk of COVID-19 among front-line health-care workers and the general community: a prospective cohort study. Lancet Public Health. 2020 Sept; 5(9). Available from: https://doi.org/10.1016/S2468-2667(20)30164-x.

6. Harris, P., Taylor, R., Thielke R., Payne, J., Gonzalez, N., and Conde, J. Research electronic data capture (REDCap)—a metadata-driven methodology and workflow process for providing translational research informatics support. J Biomed Inform. 2009 Apr; 42(2). Available from: https://doi.org/10.1016/j.jbi.2008.08.010.

7. Harris PA, Taylor R, Minor BL, Elliott V, Fernandez M, O’Neal L, Mcleod, L., Delacqua, G., Delacqua, F., Kirby, J., and Duda, S.N. The REDCap consortium: Building an international community of software platform partners. J Biomed Inform. 2019;95:103208. Epub 2019/05/09. doi: 10.1016/j.jbi.2019.103208.

8. Kataria, Y., Cole, M., Duffy, E., de la Cena, K., Schechter-Perkins, E.M., Bouton, T., Werler, M.M., Pierre, C., Ragan, E.J., Weber, S.E., Jacobson, K.R., and Andry, C. Seroprevalence of SARS-CoV-2 IgG antibodies and risk factors in health care workers at an academic medical center in Boston, Massachusetts. Sci Rep. 2021 Nov; 11(9694). Available from: https://doi.org/10.1038/s41598-021-89107-5.

9. Grupel, D., Grazit S., Schreiber, L., Nadler, V., Wolf T., Lazar, R., Supino-Rosin, L., Perez, G., Peretz, A., Ben Tov, A., Mizrahi-Reuveni, M., Chodlick, G., and Patalon, T. Kinetics of SARS-CoV-2 anti-s IgG after BNT162b2 vaccination. Vaccine. 2021 Sept; 39(38). Available from: https://doi.org/10.1016/j.vaccine.2021.08.025.

10. Feng, S., Phillips, D.J., White, T., Sayal, H., Aley, P.K., Bibi, S., Dold, C., Fuskova, M., Gilbert, S.C., Hirsch, I., Humphries, H.E., Jepson, B., Kelly, E.J., Plested, E., Shoemaker, K., Thomas, K.M., Vekemans, J., Villafana, T.L., Lambe, T., Pollard, A.J., Voysey, M., and Oxford COVID Vaccine Trial Group. Correlates of protection against symptomatic and asymptomatic SARS-CoV-2 infection. Nat Med. 2021 Sept; 2032-2040. Available from: https://doi.org/10.1038/s41591-021-01540-1.

11. Dimeglio, C., Herin, F., Martin-Blondel, G., Miedouge, M., and Izopet, J. Antibody titers and protection against a SARS-CoV-2 infection. J Inf. 20221 Sept. Available from: http://doi.org/10.1016/j.jinf.2021.09.013.

12. Guiomar, R., Santos, A.J., Melo, A.M., Costa, I., Matos, R., Rodrigues, A.P., Kislaya, I., Silva, A.S., Roque, C., Silva, C., Aguiar, J., Graca, A.S., and Machado, A. High correlation between binding IgG (anti-RBD/S) and neutralizing antibodies against SARS-CoV-2 six months after vaccination. MedRxiv [preprint] 2021 Dec. Available from: http://doi.org/10.1101/2021.12.10.21267607.

13. Levin E., Lustig, Y., Cohen, C., Fluss, R., Indenbaum, V., Amit, S., Doolman, R., Asraf, K., Mendelson, E., Ziv, A., Rubin, C., Freedhman, L., Kreiss, Y., and Regev-Yochay, G. Waning immune humoral response to BNT162b2 Covid-19 vaccine over 6 months. N Engl J Med. 2021 Dec; 385(24). Available from: https://doi.org/10.1056/NEJMoa2114583.

14. Perkmann, T., Perkmann-Nagele, N., Koller, T., Mucher, P., Radakovics, A, Marculescu R., Wolzt, M., Wagner, O.F., Binder, C.J., and Haslacher, H. Anti-spike protein assays to determine SARS-CoV-2 antibody levels: a head-to-head comparison of five quantitative assays. Microbiol Spectr. 2021 Sept; 9(1). Available from: https://doi.org/10.1128/Spectrum.00247-21.

15. Bergwerk, M., Gonen, T., Lustig, Y., Amit, S., Lipsitch, M., Cohen, C., Mandelboim, M., Levin, E.G., Rubin, C., Indenbaum, V., Tal, Illana, Zavitan, M., Zuckerman, N., Bar-Chaim, A., Kreiss, Y., and Regev-Yochay, G. Covid-19 breakthrough infections in vaccinated health care workers. N Eng J Med. 2021 Jul. Available from: http://doi.org/10.1056/NEJMoa210972.

16. Hacisuleyman, E., Hale, C., Saito, Y., Blachere, N.E., Bergh, M., Conlon, E.G., Schaefer-Babajew, D.J., DaSilva, J., Muecksch, F., Gaebler, C., Lifton, R., Nussenzweig, M.C., Hatziioannou, T., Bieniasz, P.D., Darnell, R.B. Vaccine breakthrough infections with SARS-CoV-2 variants. N Eng J Med. 2021 Apr. Available from: http://doi.org/10.1056/NEJMoa2105000.

17. Hoffmann, M., Kruger, N., Schulz, S., Cossmann, A., Rocha, C., Kempft, A., Nehlmeier, I., Graichen, L., oldenhauer, A.S., Winkler, M.S., Lier, M., Dopfer-Jablonka, A., Jack, H.M., Behrens, G.M.N., and Pohlmann, S. The Omicron variant is highly resistant against antibody-mediated neutralization: Implications for control of the COVID-19 pandemic. Cell. 2022 Feb; 185(3). Available from: http://doi.org/10.1016/j.cell.2021.12.032.

